# Experiences of care-seeking behaviour for sexually transmitted infections among gay and bisexual men: A phenomenological study

**DOI:** 10.1101/2023.02.27.23286480

**Authors:** Prince Owusu Adoma, Cecilia Adomah Yeboah Snr, Bismark Nantomah, Emmanuel Manu, Mawuli Kushitor

**Affiliations:** Department of Health Administration and Education, Faculty of Science Education, University of Education, Winneba, Ghana; Kwatire Polyclinic, Sunyani West Municipal Health Directorate, Odumase – Bono Region, Ghana; Department of Population and Reproductive Health, School of Public Health, University for Development Studies, Tamale, Ghana; Department of Population and Behavioural Sciences, School of Public Health, University of Health and Allied Sciences, Ho, Ghana; Department of Health Policy Planning and Management, School of Public Health, University of Health and Allied Sciences, Ho, Ghana

**Keywords:** Care seeking behaviour, STI, Gay and bisexual men, Bono region

## Abstract

**Introduction:** Gay and bisexual men (GBM) are stigmatized in the Ghanaian society and that negatively affect their care-seeking behaviour. We sought to understand the experiences of care-seeking behaviour (CSB) for STIs among gay and bisexual men in Bono region, Ghana.

**Methods:** A respondent-driven sampling was used to collect data from 17 gay and bisexual men in Bono region based on phenomenological qualitative approach. The data were thematically analysed using the Atlas.ti software.

**Results:** Results were presented under various themes with appropriate accompanying excerpts. Two broad themes emerged from the data; personal and health system experiences of treating STIs. Personal experiences such as economic conditions, knowledge on STI, marital experiences and bisexual’s partner awareness of sexual orientation had influences on CSB. Experience with the cost of treatment, stigmatisation by health care workers (HCW) and perceived quality healthcare were the health system factors found to influence CSB.

**Conclusion:** To help improve STIs care-seeking behaviour, government need to encourage and economically empower GBM, while at the same time, improving their knowledge on STI prevention and control. The National Health Authority should intensify and monitor the implementation of the national health insurance at the private healthcare sectors without favour and discrimination for gay and bisexual men.

## Introduction

Gay and bisexual men (GBM) are stigmatized in the Ghanaian society and that negatively affect their care-seeking behaviour. They are particularly, vulnerable to sexually transmitted infections [STIs] [1, 2]. One of the key reasons for their high vulnerability to STIs is due to the high risk of transmission of STIs through anal sex. The walls of the anus are thin and more easily torn during sexual intercourse, creating an entry point for STIs into the bloodstream [2, 3]. Low uptake of STI prevention and health services among GBM has been reported, as a result of self-reported fear of seeking health services, concerns of disclosure of sexual orientation and discrimination in healthcare settings [4]. In the Ghanaian society, the act of men having sex with men is viewed as “unnatural” and illegal [5] exacerbating its clandestine nature and stigmatisation, thereby hampering STI/HIV prevention and control among the group [6].

Prevention of STIs among GBM has longed been one of the main objectives of the National AIDS Control Programme (NACP) of the Ghana Aids Commission [7]. Moreover, the “Health For All” policy and the “Primary Health Care” strategy of the World Health Organization (WHO) [8] compels governments to provide equity of access to quality care, and also address the basic health needs of all people, including GBM [9]. To achieve this, Drop in Centers (DICs), which strive toward being free of stigma and where key populations such as Female Sex Workers and men who have sex with men (MSM) can access basic healthcare services such as HIV counseling and testing; STI screening and treatment; family planning information; and some contraceptive methods have been introduced by the Ghana government [10]. Moreover, services availability for GBM in terms of behaviour change communication, condom and lubricant distribution are also often delivered via outreach services [11].

Despite the attempt to improve health seeking behaviour for key population groups (such as GBM), data on GBM in Ghana is limited. [12]. The Ghana men’s study, conducted in 2017 in happens to be the comprehensive data on GBM. The data collected information from 4,095 MSM across 10 regions, focusing on their STI/HIV prevalence and risk behaviour [1]. Crucially, the limited information on health seeking behaviour among the GBM limits the extent to which the health system intervenes on HIV/AIDS spread within the special population. Stigma, sexual identity and a hostile environment have been implicated in non-disclosure and the secrecy associated with GBM [13, 14]. Besides, GBM is criminalized in Ghana [15, 16, 17].

The current study delved into the lived experiences of GBM in selected communities of the Bono region of Ghana, following the Standards for Reporting Qualitative Research (SROR) guidelines of qualitative reporting [18] to understand their experiences on care-seeking behaviour in relation to STI treatment and control. This will inform policy and add to the body of knowledge.

The study was pinned on the assumption that individual’s perceived threat and outcome expectation were determinants of their care seeking behaviour. The study was therefore informed by the health belief model [HBM] [19]. According to the model, health behaviour of an individual is examined through their perceptions and outcomes towards a disease or intervention. In relation to the study, GBM appraises their vulnerability of STI care-seeking, its serious, coupled with the benefits of care seeing as oppose the barriers that mitigate against care-seeking, thus influences cue to care-seeking and finally adoption of a care seeking behaviour, as depicted in Figure 1. The HBM is useful in designing interventions that can be used to increase care seeking behaviour based on patient characteristics and health system factors influencing care seeking behaviour.

**Figure 1:**
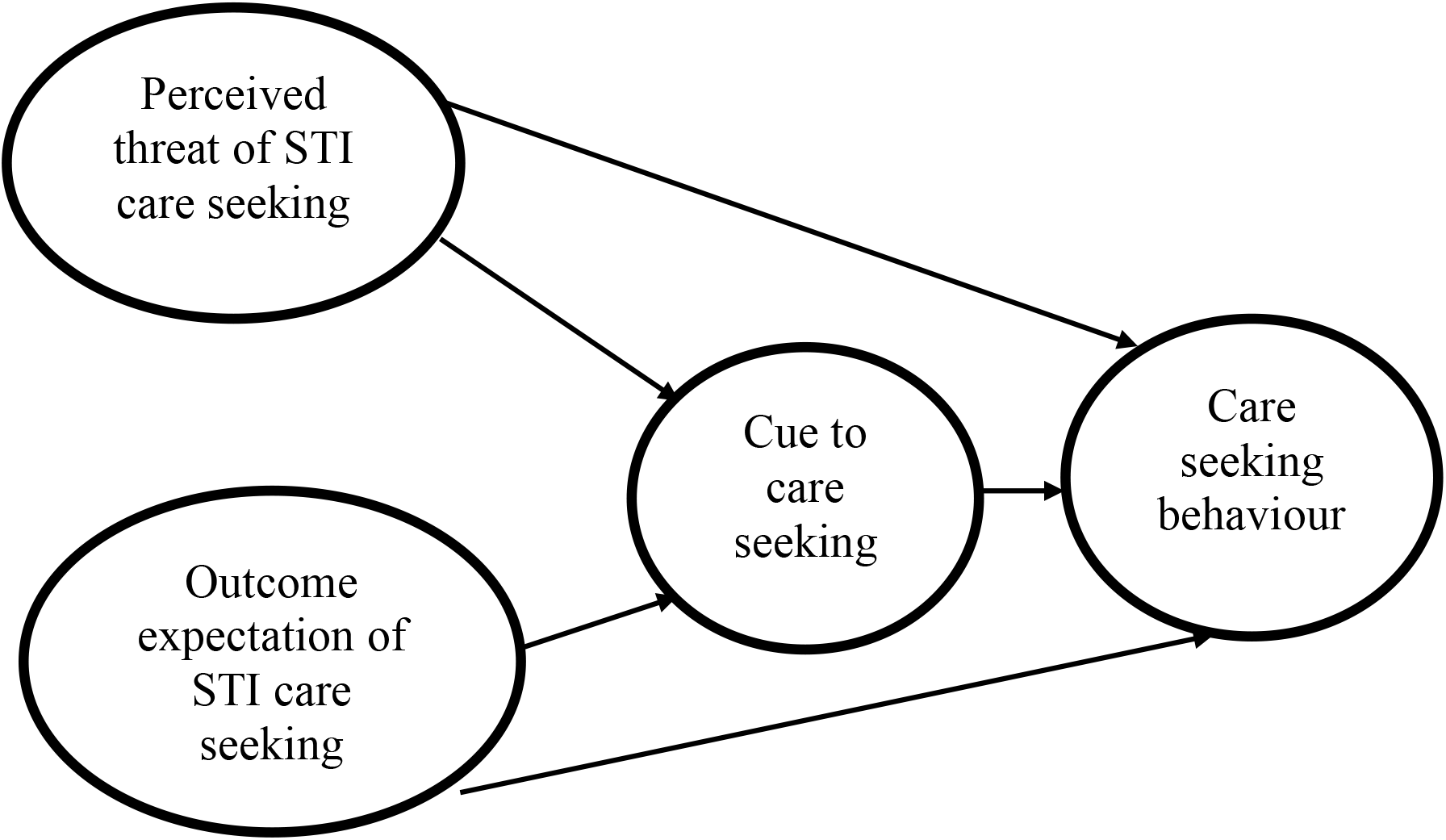
Conceptual framework for care-seeking behaviour among GBM adapted from Rosenstock [19]

## Materials and Methods

### Study Setting and Period

The study was qualitative and conducted in three selected cities (Sunyani, Berekum and Dormaa Ahenkro) in the Bono region of Ghana between April to November, 2019. The region has twelve administrative districts. The region is located in the transitional zone/middle belt of Ghana, with a population size of 1,082,508 [20]. Anecdotal evidence from the community members suggest that sex between men is common, although clandestine. This part of the country has had a long history of external migration to continental [21]. For instance, in Berekum Municipality, it is the highest migration centre in Ghana and international migration is generally considered as an integral part of livelihood and advancement strategies for most families [22] hence the choice of the region for the study.

Also, in the Bono region, only three of the districts have a DIC for GBM, reaching less than 150 GBM [1]. This suggest inadequate sexual healthcare needs among GBM in the region, creating unmet care-seeking need for STI control. However, barriers that hinder care-seeking among GBM, need to be identified and addressed so as to achieve the Sustainable Development Goal 3 which advocate for good health and wellbeing for all [23]. It is, therefore, important to understand the care-seeking behaviour of GBM in relation to STI prevention and control to reduce the STI/HIV burden among the population [24].

### Study Design

The phenomenological research design, using the key informant interview approach, was used to explore the lived experiences of health care-seeking behaviour for sexually transmitted infections among GBM in selected towns in Bono region. Phenomenology is an approach to qualitative research that focuses on the commonality of a lived experience within a particular group with the fundamental goal of arriving at a description of the nature of the particular phenomenon [25].This study, therefore, used the phenomenology to gain new insights, discover new ideas and/or increase knowledge of the experiences of care-seeking behaviour among GBM [26]. Key informant interviews were conducted to ascertain the lived experiences of GBM regarding their CSB of STIs. In order to unearth the requisite information, key informants were self-confessed GBM, with at least six months experience of engaging in the sexual behaviour.

### Segmentation Plan

The study population comprised GBM in three purposively selected communities in the Bono region of Ghana. This group of participants were selected because they carry and stand a high risk of getting STIs/HIV [27]. Thus, they carry a disproportionally high burden of HIV prevalence (18%) as compared to the general population (2%) [28]. To qualify for the study, participant had to be of sound mind and had at least six months experience of engaging in sex between men and self-identifying as GBM. Though, Kerr and Nixon [29] argue that data saturation is often reached after twelve interviews, however, in our study, the stopping criteria was informed by the theoretical saturation point.

Both purposive and snowball sampling techniques were used in the selection of communities and recruitment of participants respectively. Purposeful sampling is the intentional selection of participants based on their understanding of a phenomenon and thus, share significant and meaningful experience concerning the phenomenon under investigation [30]. Due to the sensitive nature of GBM, the snowball sampling technique was to obtain information from the targeted population. It is a method of expanding the sample by asking one participant to recommend the study to other eligible participants [31, 32]. Thus, a lead participant initially recruited by the Principal Investigator (PI) helped in the identification and recruitment of subsequent participant who also helped in the recruitment of other participants until data saturation was reached. To confirm that participants indeed were GBM, they were encouraged to recruit any known GBM apart from their sexual partners in order to avoid sample that is comprised of only men in one sexual network. Though initially hesitant, the rapport between the PI and the lead recruiter gained the trust of other participants to refer us to any known GBM. Eventually, seventeen (17) participants were recruited and interviewed.

### The Interview Guide

A semi-structured key informants’ interview guide, constructed in English and translated verbatim into the Akan language, was used to conduct the interviews. The guide was used to collect information on the socio-demographic characteristics of participants and on their experiences regarding seeking treatment for STI infections. The instrument was piloted among two known community outreach workers Bono region DIC area. These community outreach workers have similar characteristics with the GBM in study communities where the appropriateness of the interview questions in answering the research question were established [33].

### Data collection procedure

Before the onset of data collection, ethical approval was sought from ethics review Committee, Ghana Health Service. Again, all the participants were informed of the objectives and design of the study and all ethical protocols were observed. The data collection lasted for a period of six weeks. Data collection was preceded by recruitment of a lead participant who introduced the research team to potential participants. The PI then contacted and met with proposed participants in agreed secluded locations. Interview appointments were booked for participants who were willing to be interviewed at the first meeting. The interviews were conducted by the PI, under the guidance of the second and third co-authors. Recruitment and interviewing of participants continued until data saturation was met as the research process was iterative. In the end, 17 key informant interviews were conducted. Participants were given unique identifiers which aided in the identification of their demographic information and transcripts. The first participant to be interviewed was coded as P1 and the last, P17. Approximately, each interview session lasted for about an hour. All the interviews took place in locations that were safe, secure and private, for the protection of participant’s identities and eavesdropping by passer-by. The interviews were recorded with an Olympus voice recorder, after permission was sought from the participants.

### Data analysis

Recorded interviews were transcribed into the English language by a competent language editor from the University of Cape Coast, Ghana. A step-by-step thematic analysis [34] was carried out on all transcripts and the we used the Atlas.ti. software (version 8.4.15) to categorize, group and recall data. Analytical framework was guided by a process of inductive reasoning. The themes were more emergent and anticipated from the transcripts without any predetermined classification [35]. In the first step, the first, second and fourth co-authors read through the transcripts thoroughly in order to familiarise themselves with the information. The second step involved the development of emerging themes after a series of coding stages among the 3 co-authors independently. Open coding was used to identify initial codes [36] these codes were then grouped into categories according to their similarities [37]. The categories were then organized into themes. Thus, codes were combined into overarching themes that accurately depicted the data. The three analysts then compared notes, resolved discrepancies and agreed on common themes in areas where variations were found [38]. A comprehensive analysis was then made to examine the extent to which the themes contributed to an understanding of the data. All quotations for each theme were synthesized to bring out the main ideas and the most appropriate quotations used to present the descriptive results [37].

### Trustworthiness

Trustworthiness of the study findings was ensured by following several quality approaches. First, three experienced independent coders with different disciplinary backgrounds performed the analysis. Secondly, the number of quotations were observed persistently and each theme were counted to get a more accurate idea of their significance [37]. Also, discussions were held between the coders to resolve discrepancies by agreeing on appropriate themes [38]. The results were also theoretically confirmed by comparing them with existing scientific data in the discussion. The entire research process was iterative. Thus, whenever a new code was introduced, all of the transcripts were read again to ensure that the data extraction was complete and to verify that the initial classification was credible [37].

### Ethical considerations

Ethical clearance for the study was sought from the Ethics Review Committee of Ghana Health Service (GHS-REC 025/11/219). Permission was also sought from traditional rulers of the selected communities for safe community entry. Written consent was only sought from participants who were under 18 years. However, oral consent was sought from those above 18 years to protect their identity. For anonymity purposes, assent was sought from participants who were below 18 years as they did not wish their parents/guardians to know their sexual preferences. However, such participants were only allowed into the study when they were recruited by their sexual partners who were either 18 years old or older and consented for their participation. Participation was voluntary with the right to withdraw from the study at any time without repercussions. There were no direct benefits for participating in the study.

## Results

The results are presented based on the socio-demographic characteristics of the participants as well as the themes that emerged from the study.

### Socio-demographic characteristics of participants

Out of the seventeen participants interviewed for the study, most (12) of the them were in their twenties, with only one below twenty years of age and three older than thirty years. Six participants were footballers by profession, three were traders, three were students, two were unemployed and one participant refused to disclose his employment status. Thirteen of the participants were not married. With regards to educational background, nine were senior high schoolers, four were tertiary students and four completed junior high school. Most of the participants were Akans (12) by tribe, three were Ewes and two were from the Ga tribe. Nine participants disclosed that they were gay, seven were bisexual while one participant refused to disclose whether they were bisexual or gay. Eleven participants did not have children whereas most (12) starting GBM in their teens.

### Thematic results

Two broad categories (individual socio-cultural and structural factor and health system factors), with seven sub-themes, were identified to influence participants’ care-seeking behaviour towards STI treatment, based on the tents of our conceptual framework.

### Individual Socio-cultural and Structural Factor

Individual factors such as participants’ economic conditions, knowledge about STIs, marital status and awareness of sexual orientation by female partner were found to influence participants’ care-seeking behaviour towards STI infection.

### Economic status of participants

Most of the participants indicated that they faced financial difficulty as they were either students or amateur footballers who were not on regular salary. They therefore refused to seek orthodox treatment for STI due to inadequate funds. One participant explained:

> *Although I saw the symptoms of gonorrhoea early, I could not go to hospital for treatment because I had no money as at that time* (Participant 2).

This assertion was corroborated by another participant who resorted to home treatment as he did not have money to seek treatment for his suspected gonorrhoea infection at the hospital.

> *When I suspected that I had gonorrhoea, I asked a friend who had some knowledge in traditional medicine to help me treat it [gonorrhoea] because I didn’t have money to pay for the medication they will charge me at the hospital. Doctors always think that we contract the disease [STIs] through sexual intercourse, so they have the tendency of charging high if you don’t have [health] insurance cover* (Participant 13).

Financial constraints thus, affect participants’ care-seeking behaviour towards STIs, as it prevented them from seeking treatment from hospitals and resorted to the use of herbal medicine for the treatment of STIs.

### Knowledge about STIs

Another factor we identified was knowledge of participants on STIs. Participants who were knowledgeable about STIs generally took measures early enough to treat their infections. For instance, a participant who was aware that STIs are often contracted through unprotected sex, sought treatment early when they suspected signs of STI infection. He narrated:

> *Since STIs are conditions one suffer from when you have unprotected sex with an infected person, such as Gonorrhoea and HIV. Anytime I feel some tickling in my penis after I have engaged in [unprotected] sex, I immediately go to the nearest pharmacy to get medication. I know how dangerous they [STIs] can be and I also don’t want to infect my wife so I always treat myself early enough* (Participant 10).

On the other hand, participants who were less knowledgeable about STIs delayed in seeking treatment, be it traditional or orthodox treatment, as captured in the excerpts below:

> *To me STI is a disease that one can get through sexual intercourse between man and woman. I don’t do women so I’m safe and protected. Women are evil and that’s why men who sleep with them get such diseases. Since I started sleeping with men, Whenever I feel some pains in my genitals after sex, it go away without any treatment, But before [I started sleeping with men], I will have to go to the hospital before I will feel ok* (Participant 14).

Knowledge about STI infections thus play an important role in influencing care-seeking behaviour towards their treatment as it influences the perception of GBM towards STIs. While participants who were knowledgeable on STIs knew how they could negatively impact on their health, others felt they are contracted from evil women and thus, no need to be concerned about STIs since they do not have sex with women.

### Marital status of the individual

The study also found that marital status was another individual factor that influenced participants’ care-seeking behaviour regarding the treatment of STI infections. Participants who were married to the opposite sex were in haste to seek treatment whenever they felt they might have contracted STI as they did not want their spouses to suspect them of cheating.

Most of the participants were not married. It was only 4 of them who were married. Their urgency in seeking treatment is captured in the ensuing quotation:

> *I go to see my doctor any moment I realise that I could be in danger of contracting an STI. I don’t want my wife to suspect me of cheating as that could lead to a whole lot of issues. She doesn’t know I have a male sex partner so the least thing I want her to suspect is cheating but the moment I infect her with an STI, she will suspect* (Participant 10).

Thus, being married to a woman while at the same time sleeping with men put pressure on GBM to seek early treatment as infecting their female partners could lead to suspicion and eventual exposure of their sexual orientation.

### Awareness of sexual orientation by female partners

We also found that the status of almost all participants as being either gay or bisexual was unknown to their female partners. The lack of awareness of their status as gay or bisexual by their female partners, however, led to positive care-seeking attitude towards STI infections. This was so as those who were often infected sought early treatment in order prevent their female sex partners from being suspicious. A participant explained:

> *One thing you don’t want in this Community is for your female partner to be aware of your status as either gay or bisexual. I am married to a female as that is what society expect of every man but I have my reasons for sleeping (having sex) with men. That means that my status [as* bisexual*] must remain secrete so I cannot allow this small bacterial [gonorrhoea bacteria] to expose me to the whole world. The moment I keep infecting my wife, she will suspect me of infidelity so I always treat any infection I suspect to be STI as soon as I can. During those times, I travel so that my wife will not even see me taking my medication* (Participant 3).

The effort to conceal one’s sexual orientation, thus has an important influence on care-seeking towards the treatment of STIs as most GBM are in sexual relationship with women and will go all lengths to ensure that their spouses do not find out about their sexual orientation, including seeking early treatment for STI infections. This, another participant summed up by saying:

> *I cannot let a simple STI which is curable expose me to ridicule in my community. How will my wife and children feel when they realise that I am into this behaviour [*bisexual*]? It’s a terrible thing to be* bisexual *in our community* (Participant 8).

### Health system factors influencing care-seeking behaviour towards STI treatment

Aside personal factors that influenced care-seeking behaviour towards STI infections, health system factors such as inability to use the national health insurance scheme in treating STIs, fear of stigmatisation, and quality of care received offered to GBM.

### Inability to use national health insurance scheme for treatment of STIs

Participants could not use and benefit from the national health insurance scheme though their cards were valid, this is because health care providers who treat them for STIs do not accept the national insurance as a form of payment for services as captured in the narrative below:

> *I have a personal healthcare provider who doesn’t accept the health insurance for my STI treatment so I pay for my treatment. We are suffering because we are* gay, *they [private care health providers] feel we [*GBM*] are very rich and when you go to them for STI treatment, the cost is very high as compared to what ought to be paid in our healthcare system (*Participant 11).

Another participant explained:

> *Some private clinics do not accept the national health insurance so when you visit those clinics, you have to pay from your pocket. And the thing is that, due to privacy, sometimes I prefer those places* (Participant 10)

Out of pocket payments due to non-acceptance of the national health insurance cover as form of payment for treatment of STIs hampered positive care-seeking behaviour towards treatment as private healthcare providers take advantage of the status of GBM and exploit them financially.

### Quality of healthcare received

The quality of care offered to GBM during healthcare visits played a major role in influencing their attitude towards STI treatment. This is so because although some Physicians already knew the sexual orientation of their clients, they did not judge them or discriminated against them. This positively impacted on the care-seeking behaviour of GBM towards STI treatment. A participant explained:

> *As for Mr… [A nurse], he’s good and very patient with me. Although his charges are high he will take care of me any time I go for treatment [for STI infection) even without money and I will pay him later (*Participant 5).

Another participant shared his experience on quality of care he receives from healthcare providers.

> *The doctor I go to is very good and secretive. Although he knows my status [*GBM*], he has kept it to himself. He even knows my wife but will never tell her [of my status]. Oh, and he is also gentle and caring. I even wished he could be my partner [laughs]* (Participant 13).

Quality healthcare provision was seen to have a positive influence on care seeking behaviour towards STI treatment. The more participants trusted a healthcare provider, the likelihood of them seeking treatment for STI infections.

### Stigmatisation and discrimination by healthcare providers

Most of the participants indicated that sometimes they visit their trusted healthcare providers at their residence for STI treatment despite it costing them more than they would have paid at a public hospital as these care providers already know their sexual orientation. This is because GBM are often discriminated against by healthcare providers in various public health facilities when they get to know their sexual orientation and at times disclose their status to colleagues and the general public, leading to stigmatisation and discrimination. One of the participants explained:

> *…nothing will compel me to go to the government hospital while I can privately seek for care without any trouble and avoid unnecessary questions. Nurses in our [public] hospitals talk a lot and once they know your status [as* gay*], they will spread it among themselves and even in the community. I don’t trust them to go to them for treatment (*Participant 9)

Explaining how in explaining how his status became known in a community he previously lived in, a participant mentioned:

> *It was a nurse that revealed my status to a female friend of mine some time ago at where I used to live previously. She [the nurse] told her [his friend] everything about me when I went to their facility with some sores in my anus and told them what happened to me. Since then, I only visit private Doctor* (Participant 14).

Disclosure of participant’s status by health workers in public health facilities leading to discrimination thus affect care-seeking for the treatment of STIs among GBM.

## Discussion

This study sought to understand the care-seeking behaviour for sexually transmitted infections among GBM in the Bono Region of Ghana. Two key factors; individual and health system related factors, were found to influence the care-seeking behaviour of participants for STIs. Individual factors that influenced care-seeking behaviour included economic condition of a participant, knowledge about STIs, marital status of participants and awareness of gay or bisexual status by female partner in the case of those who were bisexuals.

With reference to how economic status influenced care-seeking behaviour for the treatment of STIs, participants indicated that being a student or an amateur footballer meant they did not receive regular income. This, they explained led them to resort to herbal treatment for STIs, as orthodox treatment in public hospitals was expensive. Socio-economic status has been reported in the literature as a barrier to care-seeking for a number of health conditions [39-41]. For instance, when faced with economic crisis, most patients prefer a spiritual or traditional healer as they charge moderate prices [41]. This was the case for some participants in this study. As a result of the high cost of treatment for STIs for non-NHIS subscribers, they were pushed to resort to herbal treatment. Although the popularity of herbal medicine is high in Ghana, the efficacy and toxicity of some products are empirically unascertained and could jeopardise the health condition of those that patronise them [42]. This could lead to chronic STI infections among GBM as they resort to herbal treatment. Knowledge on STIs was another factor that the study found to influence care-seeking behaviour among participants for the treatment of STIs. Participants who were aware of the types, symptoms and severity of STI infections sought early treatment while those who were less knowledgeable attributed STI infections to superstition. Thompson and colleagues [43] concluded that knowledge on illness prevention influence care seeking among patients. When the cause, severity and prevention of an illness are poorly understood, patients delay in seeking care as they downplay the long-term effects of the disease on their health [44]. This was the case with some participants in the current study. The implication of this finding is that GBM with poor knowledge on STIs are likely to delay their treatment or poorly treat their infections, leading to a chain of transmission among GBM and the general population as some participants were bisexuals. Effective education on STIs among GBM is therefore required in the study communities and the Bono region at large in order to control the spread of STI infections among GBM and the general population.

Moreover, marital status and awareness of one’s status as being gay or bisexual were other individual factors that influenced care-seeking for STI treatment. These factors were interlinked as participants who were married to females, hence were bisexuals, sought early treatment for suspected STI infections. In some cases, they did travel to different locations to avoid being queried by their female partners. By implication, they did not want their female partners to be aware of their gay or bisexual status and would go every length to conceal their true sexual orientation. In Ghanaian settings, being labelled as gay or bisexual is a shameful sexual behaviour leading to widespread stigmatisation and societal ridicule [45]. Due to societal pressure as a result of culturally stereotyped gender roles, most GBM are married to women [46] Open disclosure of one’s gay or bisexual status could therefore shatter their marital lives. Hence, for participants married to the opposite sex, care-seeking behaviour for STI prevention and treatment is paramount. The practice of marrying female partners should therefore be encouraged in the GBM community as it serves as a self-check for prompt STI care-seeking and thus leads to improved sexual health among GBM.

With reference to health system factors, inability to use the national health insurance scheme in treating STIs, fear of stigmatisation by healthcare providers, and quality of care received offered to GBM at various health centres influenced care-seeking behaviour for STI treatment among GBM. Participants who sought care for treatment of STIs from healthcare providers disclosed their inability to use the national health insurance card to pay for STI-related services in such facilities as a barrier to poor care-seeking behaviour for STI treatment. Ghana officially launched the national health insurance scheme in August 2003 [47] and was envisaged to cover the cost of treatment for most ailments, including STIs. In fact, the national health insurance scheme covers the cost of treatment for all STIs, including HIV in all government owned health facilities as well as accredited private health care facilities across the country [48]. However, there are few private health centres that have not been certified by the national health insurance scheme. Hence, seeking treatment for STIs in some of such unaccredited health facilities may incur out-of-pocket payment. In reference to most of the study participants who were not gainfully employed while preferring private healthcare, they are bound to have poor care-seeking for STIs as evidenced in the transcripts. GBM in the Bono region would therefore be educated on private healthcare centres that are accredited by the national health insurance scheme to ensure that they seek care for the treatment of STIs while their privacy is protected as that is the ultimate reason for seeking care from private healthcare providers [49].

Moreover, the quality of care received by GBM in seeking care for the treatment of STIs influenced their care seeking behaviour. In instances where the healthcare provider was caring, secretive or both, participants were more than willing to report STIs for treatment. Quality healthcare has been found to have positive impact on care seeking behaviour in many settings [50, 51] and is particularly so for all, irrespective of one’s sexual orientation [52]. However, owing to the stigma associated with GBM, healthcare providers, when aware of the sexual orientation of GBM, need to be a bit sensitive in dealing with their GBM clients as any sign of discrimination or perceived poor care could lead to non-utilisation of health services. In this regard, there is the need of training healthcare professionals in Ghana to take into consideration the healthcare needs of GBM, especially in cases of STI infections.

Furthermore, fear of stigmatisation by healthcare providers served as a barrier to seeking care for STI infections among some participants. Some participants had bad experiences from health workers whereby their status as GBM were disclosed to the public thereby leading them to stigmatisation and discrimination. Healthcare providers’ discrimination of GBM has been extensively reported in the literature [53-55]. However, the practice continues, especially in Africa [56] where gender roles and aspirations make it difficult for even healthcare providers to come to terms with the practice of GBM. Moreover, in Ghana, stigmatisation and discrimination by some health workers often occur in public health centres than private ones [57] discouraging the use of public healthcare facilities among our study participants. It is therefore important to state that healthcare providers in the Bono region and in Ghana as a whole need reorientation in order to become culturally adaptive to work with people from varied cultural and sexual preferences, including GBM.

### Strengths and limitations of the study

The strengths of the current study are that, the true status of participants as GBM was ascertained by ensuring participants recruited their sexual partners into the study. Moreover, the inductive data analysis process ensured that the results were not influenced by the researchers’ pre-conceived assumptions. One of the limitations of the study is that, participants were selected from only three communities in the region. Hence, the smaller number of sampled communities and the total sample size makes it difficult for generalisation of the results on similar participants from different parts of the region and the country at large. The results should therefore be interpreted with caution. Despite this limitation, the results present a fair idea on care seeking behaviour of GBM in relation to STI treatment and could therefore be used as basis for further research.

## Conclusion

Care-seeking behaviour for the treatment of STIs was found to be influenced by seven factors; four individual and three health-system related factors. To help improve care-seeking behaviour for the treatment of STIs among GBM in the bono region, there is the need to economically empower those indulged in GBM behaviour while at the same time improving their knowledge on STI infections. Lastly, private healthcare providers should be encouraged to accept active NHIS cards as a form of payment for STIs in order to avoid out of pocket payments for GBM who utilise private health facilities for the treatment of STIs.

## Data Availability

All data produced in the present study are available upon reasonable request to the authors

## Acknowledgements

We acknowledge our participants and community leaders of the selected communities where this study was carried out for their contributions in helping to make the publication of this manuscript a reality.

## Availability of data and material

Data will only be provided upon **reasonable** request and for **only** academic purposes.

## Author Contributions

Conceived and designed the study: CAY and POA. Analyzed the data: POA, CAY and EM. Wrote the paper: POA, EM, BN, CAY and MK. Reviewed the available literature and performed the analyses: POA, CAY, BN, EM and MK. Contributed to the interpretation of results and write up: POA, EM, BN and MK.

